# Detection of Lung Cancer Cases in Chest CT Scans Utilizing Artificial Intelligence: A Retrospective Analysis of Data During the COVID-19 Pandemic

**DOI:** 10.1101/2023.12.26.23299170

**Authors:** R.A Zukov, I.P. Safontsev, M.P Klimenok, T.E. Zabrodskaya, N.A. Merkulova, V.Yu Chernina, M.G. Belyaev, M.Yu Goncharov, V.V. Omelyanovsky, K.A. Ulyanova, E.A. Soboleva, M.A Donskova, M.E. Blokhina, E.A Nalivkina, V.A Gombolevskiy

**Affiliations:** Federal State Budgetary Educational Institution of Higher Education «Prof. V.F. Voino-Yasenetsky Krasnoyarsk State Medical University» of the Ministry of Healthcare of the Russian Federation (FSBEI HE Prof. V.F. Voino-Yasenetsky KrasSMU MOH Russia), 1 Partizana Zheleznyaka Street, Krasnoyarsk, Russia, 660022; Krasnoyarsk Regional Clinical Oncological Dispensary named after A.I. Kryzhanovskogo, (KRCOD named after A.I. Kryzhanovskogo) Krasnoyarsk, Russia, 660133, 16, 1st Smolenskaya St.; IRA Labs, Moscow, Russia, Skolkovo Innovation Centre territory, 30, Bolshoi boulevard, bld. 1.; Artificial Intelligence Research Institute (AIRI), Moscow, Russia, 32 Kutuzovsky Prospekt 1, President Plaza Business Centre, Moscow; Skolkovo Institute of Science and Technology Autonomous Non-Profit Educational Organisation of Higher Education, Moscow, Bolshoy Boulevard, 30, Bld. 30, Bld. 1.; Center for Expertise and Quality Control of Medical Care, Moscow, Russia; Federal State Budgetary Educational Institution of Further Professional Education “Russian Medical Academy of Continuous Professional Education” of the Ministry of Healthcare of the Russian Federation (FSBEI FPE RMACPE MOH Russia), Moscow, Russia; Federal State Budgetary Institution “Financial Research Institute” (FSEI Financial Research Institute), Moscow, Russia; Russian Research of Health (RIH), Moscow, Russia; Institute for Information Transmission Problems of the Russian Academy of Sciences (Kharkevich Institute), Moscow, Russia; AstraZeneca Pharmaceuticals LLC, Moscow, Russia; World-Class Research Center “Digital biodesign and personalized healthcare”, Sechenov First Moscow State Medical University, Moscow, Russia

**Keywords:** Lung cancer, computed tomography, artificial intelligence, chest, health economics, performance measurement

## Abstract

**Purpose:** To evaluate the potential of using artificial intelligence (AI) focused pulmonary nodule search on chest CT data obtained during the COVID-19 pandemic to identify lung cancer (LC) patients.

**Methods:** A multicenter, retrospective study in the Krasnoyarsk region, Russia analyzed CTs of COVID-19 patients using the automated algorithm, Chest-IRA by IRA Labs. Pulmonary nodules larger than 100 mm³ were identified by the AI and assessed by four radiologists, who categorized them into three groups: “high probability of LC”, “insufficiently convincing evidence of LC”, “without evidence of LC”. Patients with findings were analyzed by radiologists, checked with the state cancer registry and electronic medical records. Patients with confirmed findings that were not available in the cancer registry were invited for chest CT and verification was performed according to the decision of the medical consilium. The study also estimated the economic impact of the AI by considering labor costs and savings on treatment for patients in the early stages compared to late stages, taking into account the saved life years and their potential contribution to the gross regional product.

**Results:** An AI identified lung nodules in 484 out of 10,500 chest CTs. Of the 484, 355 could be evaluated, the remaining 129 had de-anonymization problems and were excluded. Of the 355, 252 cases having high and intermediate probabilities of LC, 103 were found to be false positives. From 252 was 100 histologically verified LC cases, 35 were in stages I-II and 65 were in stages III-IV. 2 lung cancers were diagnosed for the first time. Using AI instead of CT review by radiologists will save 2.43 million rubles (23,786 EUR/ 26690 USD/ 196,536 CNY) in direct salary, with expected savings to the regional budget of 8.22 million rubles (80,463 EUR/ 90,466 USD/ 666,162 CNY). The financial equivalent of the life years saved was 173.25 million rubles (1,695,892 EUR/ 1905750 USD/ 14,033,250 CNY). The total effect over five years is estimated at 183.9 million rubles (1,800,142 EUR/ 2,022,907 USD/ 14,895,949 CNY).

**Conclusion:** Using AI to evaluate large volumes of chest CTs done for reasons unrelated to lung cancer screening may facilitate early and cost-effective detection of incidental pulmonary nodules that might otherwise be missed.

## Introduction

Lung cancer (LC) remains a leading cause of cancer morbidity and mortality worldwide, with 2.2 million cases and 1.8 million deaths reported annually [1]. In the Russian Federation (RF), the standardized incidence rate of lung cancer is 20.8 per 100,000 population, representing a 2.7% increase compared to 2020 [2]. Krasnoyarsk Krai, a region within the RF, has experienced an unfavorable epidemiological situation regarding lung cancer, ranking third in cancer morbidity after breast and skin cancer.

Notably, since 2020, the COVID-19 pandemic has led to a significant increase in the number of chest computed tomography (CT) scans performed in Krasnoyarsk Krai (36,577 in 2019 and 236,234 in 2021). However, despite the increased imaging, there has been a 5.2% decrease in lung cancer incidence in the region compared to 2019.

A potential explanation for the current predicament is the overlooked presence of nodular lung masses amidst the characteristic alterations associated with COVID-19 viral pneumonia. Consequently, there is a demand for novel diagnostic techniques designed to enhance the efficacy of detecting radiologically occult LC. One such approach involves the incorporation of artificial intelligence (AI) as a component of a hybrid diagnostic strategy for LC. In this two-stage process, an AI system initially screens large volume CTs, followed by expert radiologist review of the AI-selected cases to make a final determination. In 2022, the Krasnoyarsk region participated in the pilot project “Retrospective Analysis of CT using AI Algorithm Chest-IRA from IRA Labs.” The available literature did not find studies that examined the detection of new lung cancers through retrospective application of AI on CT data collected during the COVID-19 pandemic.

The objective of this investigation is to evaluate the feasibility of employing the AI algorithm to identify patients with a high likelihood of LC based on chest CT data acquired during the COVID-19 pandemic.

## Materials and Methods

This study was an observational, multicenter, retrospective investigation. Ethical committee approval was obtained, and informed consent was waived.

### Inclusion Criteria

- adult patients (age ≥ 18 years) from the Krasnoyarsk Krai region;
- patients seeking medical care at primary and specialized healthcare facilities;
- CTs performed for the evaluation of COVID-19 infection;
- CTs performed in the period between November 1, 2020, and February 28, 2021;
- CTs performed without intravenous contrast;
- availability of CT images in DICOM format.

### Exclusion Criteria

- absence of CT images in DICOM format;
- false-positive errors in AI detection;
- CT slice thickness exceeding 1.5 mm;
- incomplete chest CT scan data acquisition;
- presence of significant motion artifacts;
- inability to retrieve patient data post-anonymization.

### CT scanners

CTs were performed on Toshiba Aquilion 64, GE Revolution EVO and GE Healthcare Revolution Discovery CT scanners using a standard protocol: tube voltage 120 kV, with automatically adjusted X-ray tube current strength, tube rotation speed (Time rotation) 0.50 s, pitch 0.938, slice thickness 1 mm.

### Data security

The study was conducted in compliance with all legislative requirements concerning personal data, as data transmission was performed after preliminary anonymization and assignment of a unique encryption key to each investigation. This allowed for the subsequent identification of patients requiring further examination (with lung lesion characteristics) at the oncology dispensary. The data were depersonalized and directed to the platform of “IRA Labs.”

### AI algorithm

All included CT studies were processed using the AI algorithm “Chest-IRA” (version LungNodules-IRA, v2.4). It is one of the algorithms of a complex AI service that is capable searching for 11 different pathologies in chest CT [3]. This AI algorithm was validated on a specially prepared calibration dataset within the framework of the Moscow Experiment on the application of AI [3].

According to methodological recommendations for clinical trials of software based on intelligent technologies, the “Chest-IRA” algorithm was approved for use, as it exceeded the threshold accuracy value of no less than the receiver operating characteristic area under the curve (ROC AUC) of 0.81 [4]. The values of independent AI developer evaluations of diagnostic accuracy metrics for the “Chest-IRA” algorithm were as follows: ROC AUC 0.96; sensitivity 0.94; specificity 0.94; accuracy 0.94 [3].

This AI algorithm is based on two neural networks. The first neural network (H1) addresses the problem of semantic segmentation of pulmonary nodules and masses. H1 was trained on the LIDC-IDRI dataset [5], which is publicly available and includes CT studies for 1,018 patients with lung nodule partitioning in the form of binary masks. H1 produces a binary mask for the input image, which is then decomposed into connected components. Some of these components correspond to pulmonary nodules and masses, while the rest are false positives. A second neural network (H2) tackles the problem of classifying the connected components into true positives and false positives to reduce the number of false positive findings. H2 was trained on a dataset consisting of 2,351 CT studies using public data with markup in the form of coordinates of different H1 findings with classification labels.

The IRA Labs algorithm has established itself as the first capable of making a differential diagnosis as a triage study between CT scans with the presence of histologically confirmed LC, bacterial pneumonia, viral pneumonia characteristic of COVID-19, or normal findings [6].

The Chest-IRA algorithm (LungNodules-IRA, v2.4) was employed to analyze images, identifying lung nodules with a volume exceeding 100 mm³ and providing their localization, size, and volume. The algorithm was designed to exclude calcifications and not identify them as nodular masses. However, the algorithm had several limitations:

1. The study must include at least one axial series that encompasses the lungs.
2. The distances between the slices in this series must be equal for more than 95% of thes lices and should not be less than 3 mm.
3. The series must cover an area larger than 192mm x 192mm x 96mm.

### Data workflow

CT-exams with AI-detected findings were reviewed by four radiologists from the Krasnoyarsk Krai State Budgetary HealthCare Institution “Krasnoyarsk Krai Clinical Oncological Dispensary named after A.I. Kryzhanovsky” (KKCOD), each possessing over 10 years of experience in thoracic radiology. Each study was reviewed once by a single radiologist. During image analysis, radiologists classified the images into three categories: “patients with high probability of radiologic LC” (CT signs indicative of malignant masses), “patients with no evidence of LC” (false positive algorithm triggers), and “patients with insufficiently convincing evidence of LC”. The high probability group included patients with solid nodules of 6 mm or larger in diameter (or more than 100 mm³) and more than five foci exhibiting benign characteristics as Guidelines for Management of Incidental Pulmonary Nodules by the Fleischner Society [7]. Nodules with “ground-glass” appearance smaller than 6 mm in diameter (or more than 100 mm³), perifissural nodules, and benign calcifications were considered as signs of benign masses. Studies that did not meet the aforementioned criteria were classified under the category of “patients with insufficiently compelling evidence of LC”.

In each case, patient medical records and CTs underwent examination to establish subsequent strategies by the radiologists. If the radiologist confirmed the pulmonary nodule suggested by the artificial intelligence algorithm, each such patient was checked into the cancer registry and the mortality registry.

### Patient workflow

If the patient was not on the cancer registry and was not listed as deceased, the patient was invited for a follow-up chest CT scan to evaluate the dynamics of the pulmonary nodule that was detected by the AI and confirmed by the radiologist. Based on the results of the follow-up CT study, the medical consilium made a decision on the management tactics for each patient, including hospitalization with biopsy for histological verification or surgery.

### Economics

To assess the economic impact of implementing the AI algorithm, labor expenses were determined by considering the mean salary of radiologists in Krasnoyarsk Krai. Furthermore, the cost savings associated with early-stage LC treatment compared to late-stage interventions were taken into account [8]. Additionally, an estimation method was employed to evaluate the economic effect, factoring in the saved life years and their potential contribution to the gross regional product. All detected lung cancers using AI were included in the calculation.

### Statistical analysis

Descriptive statistical methods were used in the statistical analysis.

## Results

A total of 10,500 CTs, with a male-to-female ratio of 56% to 44% and an age range of 28 to 91 years, were obtained from the regional Picture Archiving and Communication System (PACS) archive, fulfilling the established criteria (Fig.1).

**Figure 1.**
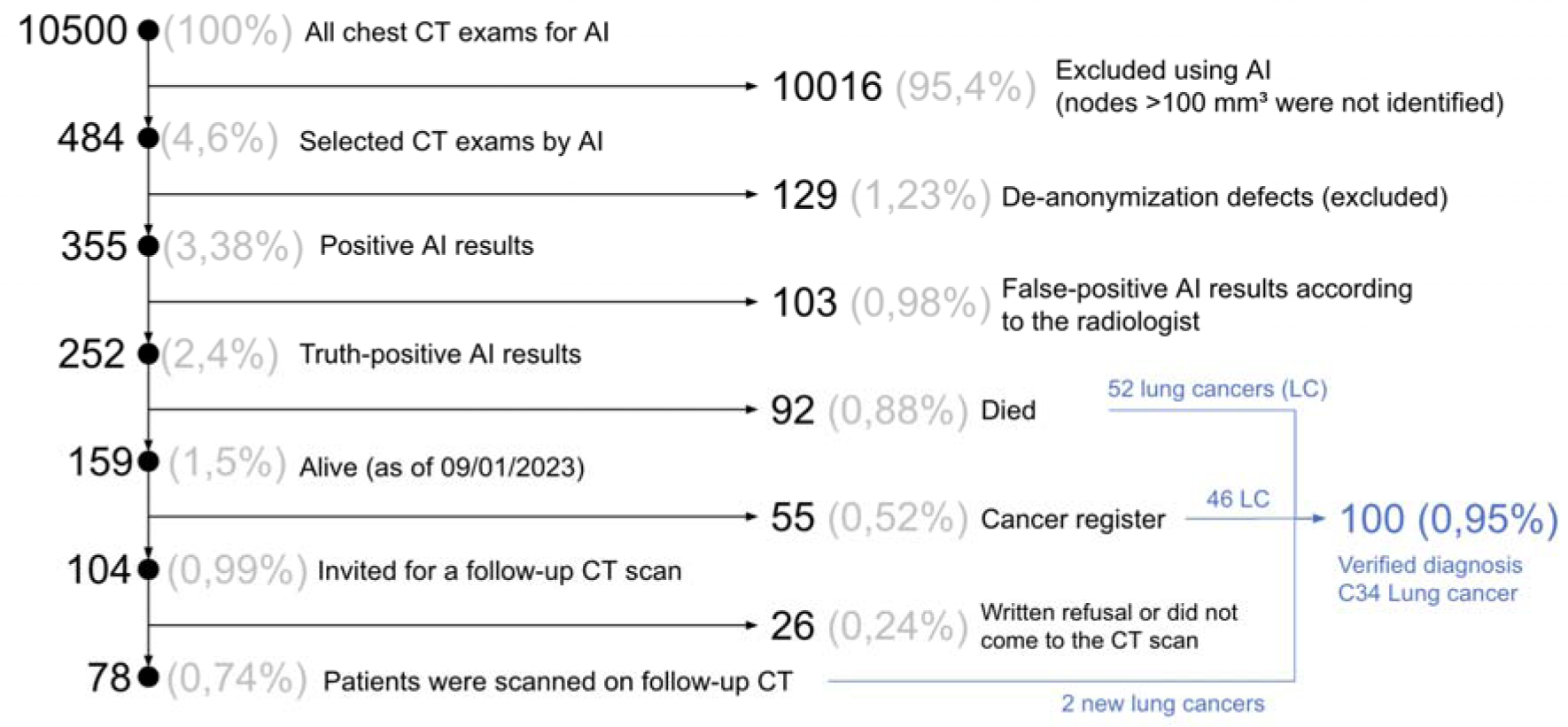
The flowchart of study.

In the pilot study, the AI algorithm identified LC indications in 484 CTs, representing 4.6% of the cases. Due to issues encountered during the personalization process, 129 CTs (26.6%) were excluded from the analysis. Further work was performed on data from 355 CTs (3,4%) only.

Upon 355 CTs of the studies selected by the AI algorithm, radiologists classified the CTs into three distinct categories (Fig.2):

1. Patients with a high probability of LC: This group comprised 192 CTs, accounting for 54% of the total studies identified by the AI (1.83% of all studies).
2. Patients with no evidence of LC: This category included 103 CTs, representing 29% of the studies identified by the AI (0.98% of all studies).
3. Patients with insufficient evidence of LC: This group consisted of 60 CTs, making up 17% of the studies identified by the AI (0.57% of all studies).

**Figure 2.**
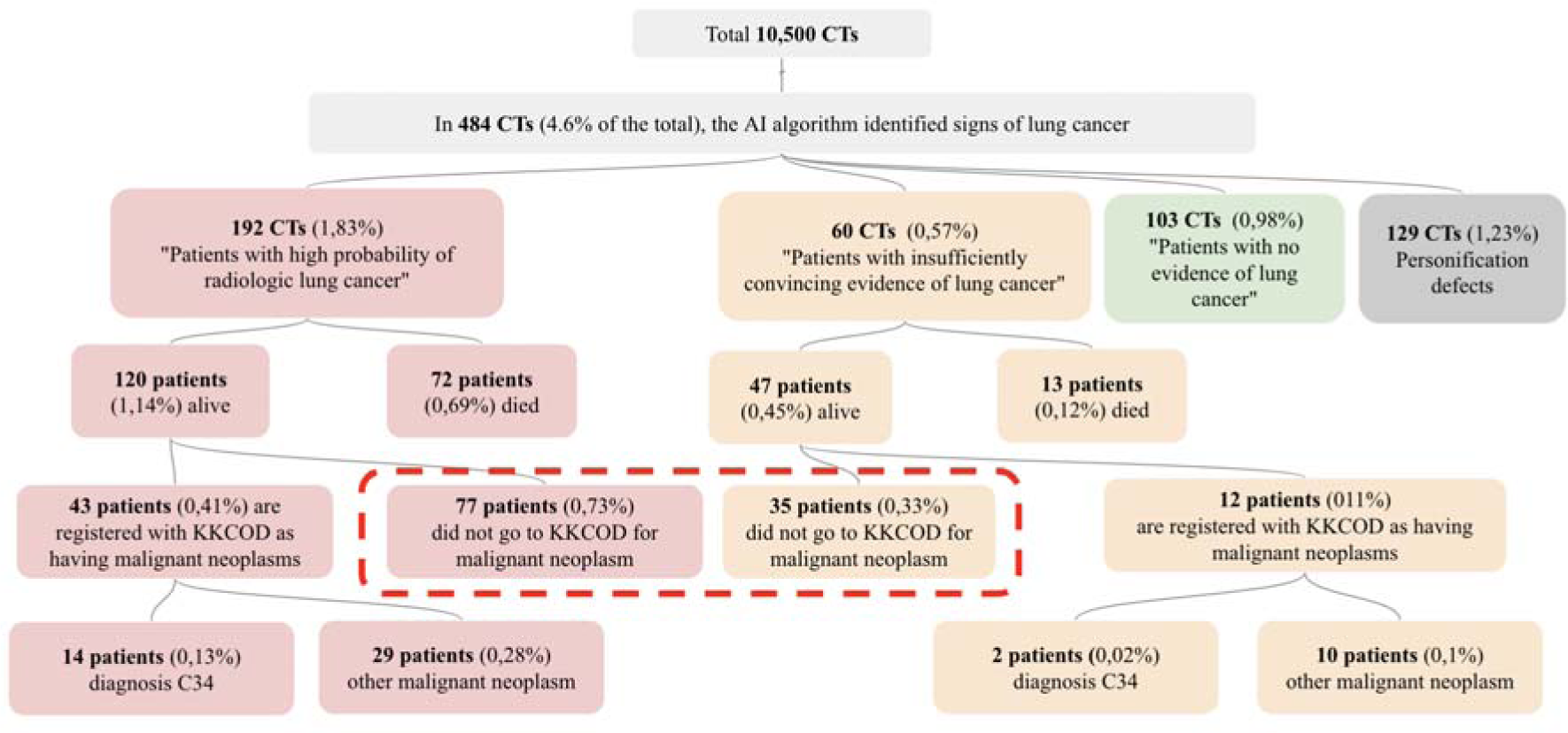
Quantitative study results. Out of the 134 patients identified (as represented by the dotted line), none sought medical treatment for their diagnosed malignant disease. Among these patients, 104 were subsequently invited to undergo a repeat chest CT scan.

The results of the AI algorithm are shown in Figure 3 and Figure 4. The time interval between CT scanning and AI analysis is from 762 to 881 days.

**Figure 3.**
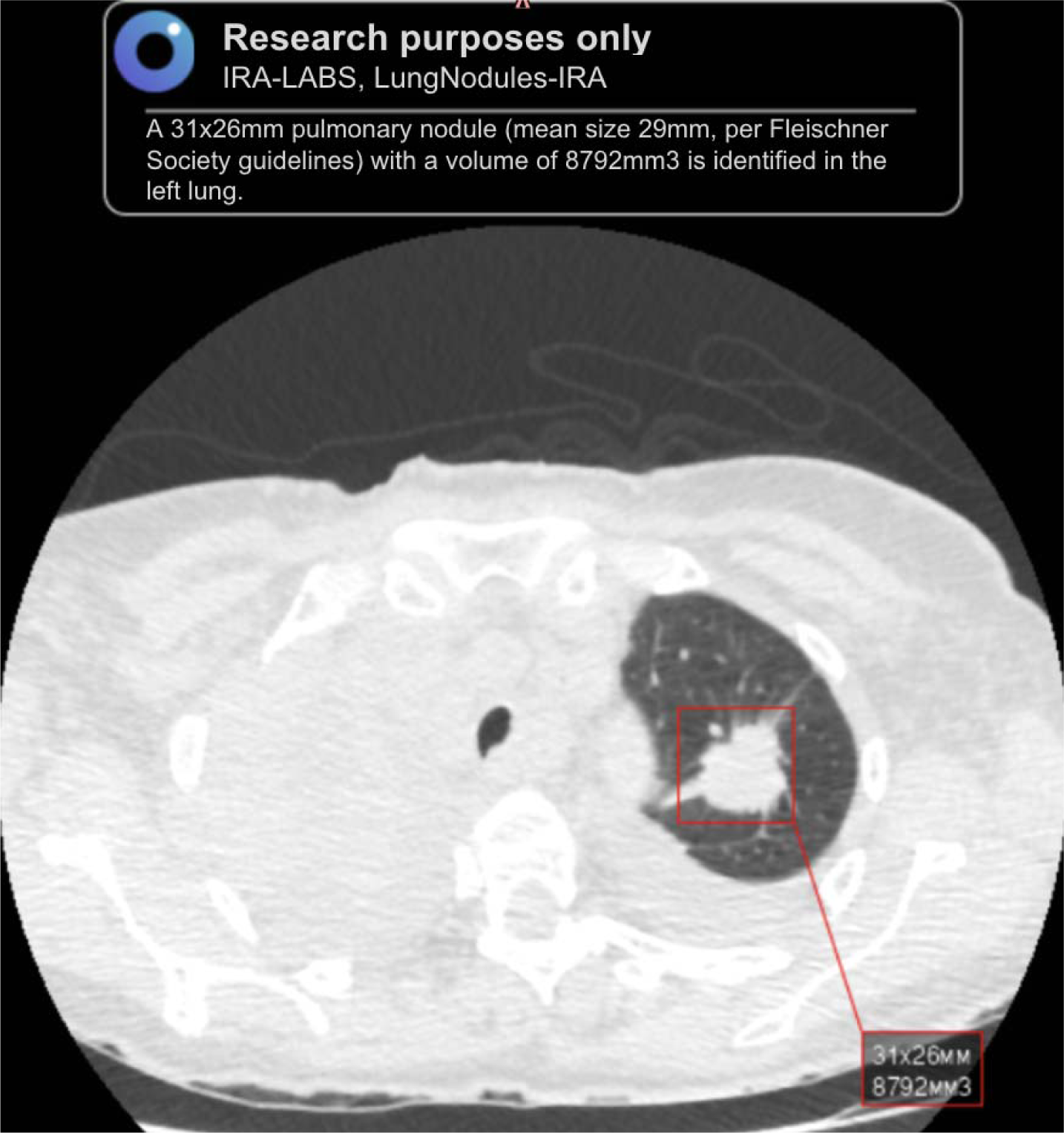
The AI algorithm labeled the pulmonary nodule in the left lung with a red square. The radiologist identified this study in the “patients with high probability of radiologic lung cancer” group at review.

**Figure 4.**
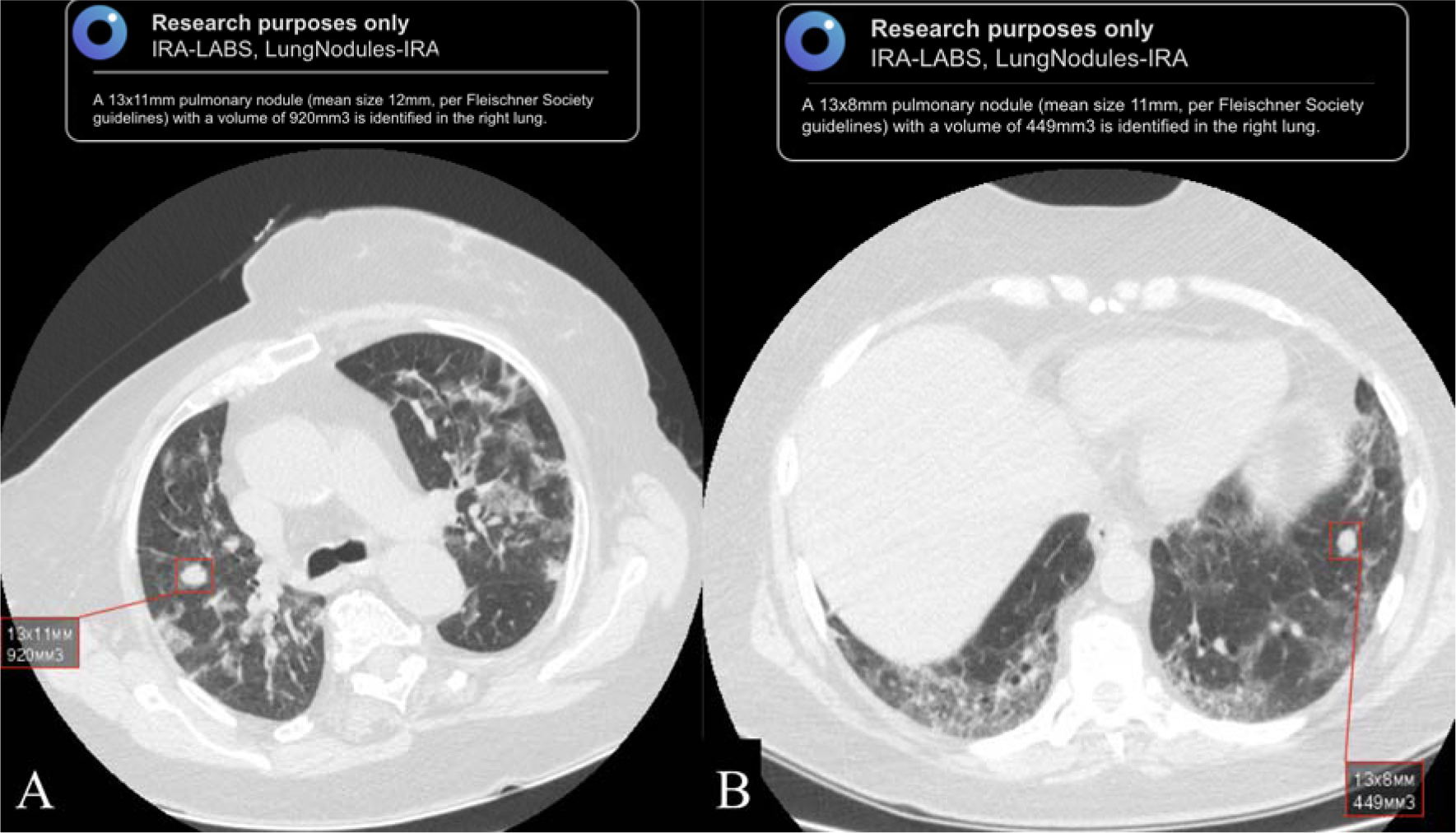
The AI algorithm labeled pulmonary nodules with red squares in the right (A) and left (B) lungs. The radiologist identified both studies in the “patients with insufficiently convincing evidence of lung cancer” group on review.

Out of 252 patients categorized into the groups “patients with a high likelihood of LC” and “patients with insufficiently compelling evidence of LC” as of January 9, 2023:

- 93 patients have passed away. Among these, 52 patients were under observation for lung and bronchial malignancies and other cancers with lung metastases at the time of the study. 11 patients were being monitored for other cancers without lung metastases, and 30 patients were not under observation for any form of cancer in the cancer registry.
- 159 patients are still alive. Of these, 55 patients were registered with lung and bronchial malignancies at the time of the study in the cancer registry. This includes 46 patients with malignant lung and bronchial neoplasms and other malignant neoplastic diseases (MND) with lung metastases (17 of these patients had malignant lung and bronchial neoplasms: 10 patients were at stage I-II, and 7 patients were at stages III-IV). 9 patients had other MNDs without lung metastases.
- 104 patients were not receiving treatment for malignant diseases (not included in the cancer registry) and were invited for a repeat chest CT. Of these, 9 patients formally declined further examination and treatment. 17 patients did not attend their follow-up examination. 78 (75% of 104) patients underwent examination and histologic verification: 2 patients were first diagnosed with non small cell LC (stages Ia and Ib) (Fig.5), and 76 patients were diagnosed with other lung pathologies (Table 1).

**Figure 5.**
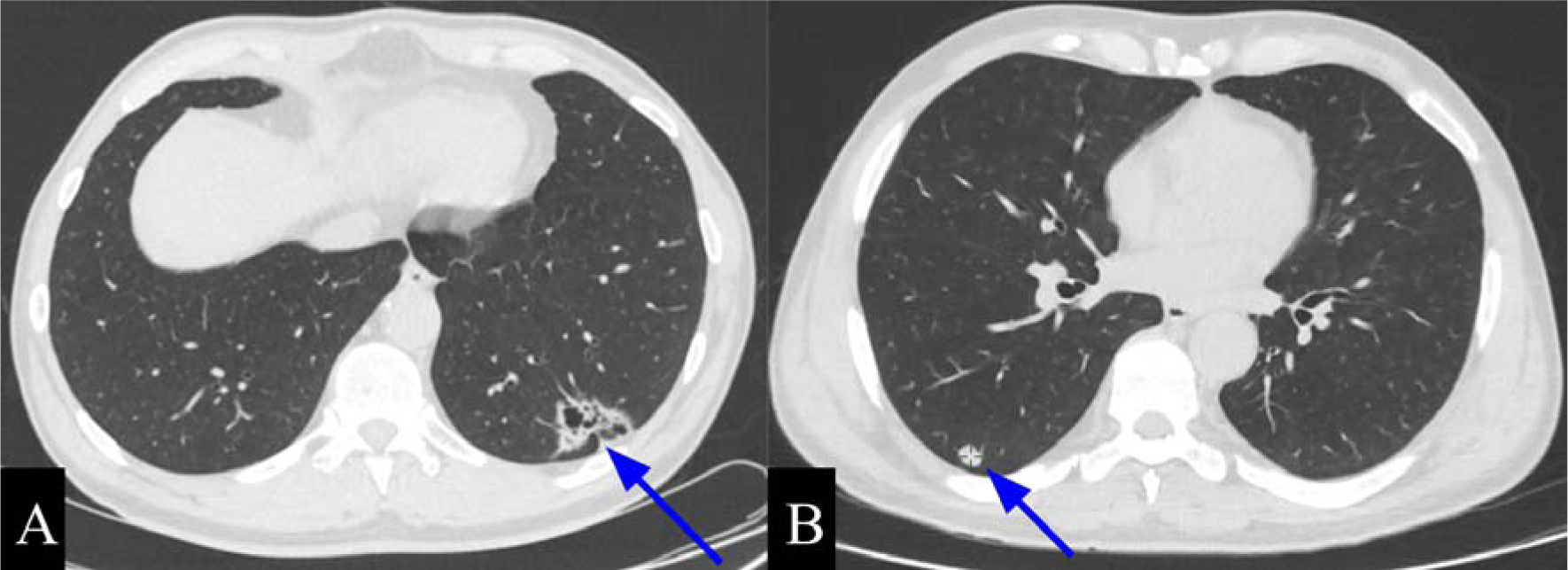
Chest CT scans of patients with confirmed lung cancer (indicated by blue arrows): A, cystic-solid mass in the left lung (stage Ia); B, solid mass in the right lung (stage Ib).

**Table 1.** Distribution of identified pathological changes in the lungs of patients who underwent a follow-up examination.

| Pathology | Number of patients |
| --- | --- |
| Hamartomas | 17 |
| Lung nodules that have reduced in size since 2020 | 13 |
| Post-inflammatory changes | 12 |
| Tuberculomas | 7 |
| Benign tumors of the lung and bronchus (D14.3) | 2 patients - surgical intervention at KKCOD, 2 patients - consultation at KKCOD |
| Sarcoidosis | 3 |
| Postoperative changes | 2 |
| Cysts | 2 |
| Fibrotic changes | 1 |
| Chronic obstructive pulmonary disease | 1 |
| Pneumosclerosis | 1 |
| Other types of interstitial lung diseases that involve fibrosis (J84.1) | 1 (surgery in the oncodispensary of St. Petersburg) |
| No Pathology | 12 |

In conclusion, out of 355 cases analyzed, 100 (28.2%) cases of LC were confirmed by AI. 2 lung cancers were diagnosed for the first time. It is noteworthy that AI also facilitated the detection of other lung pathologies. The proportion of LC cases detected at stages I-II was 35%, while those detected at stages III-IV was 65%.

An analysis of CTs, which were classified by specialists as “without signs of LC,” revealed that in the majority of cases (71 patients, 68.9%), the AI algorithm identified fibrotic changes in lung tissue. In 14 patients (13.6%), the AI algorithm marked areas of infiltration, while in 11 patients (10.7%), it highlighted hamartomas. Additionally, the AI algorithm detected pulmonary tissue vessels in 3 CTs (Fig. 6). In a few instances, the AI algorithm also labeled lymph nodes (1.0%) and tuberculomas (1.0%).

**Figure 6.**
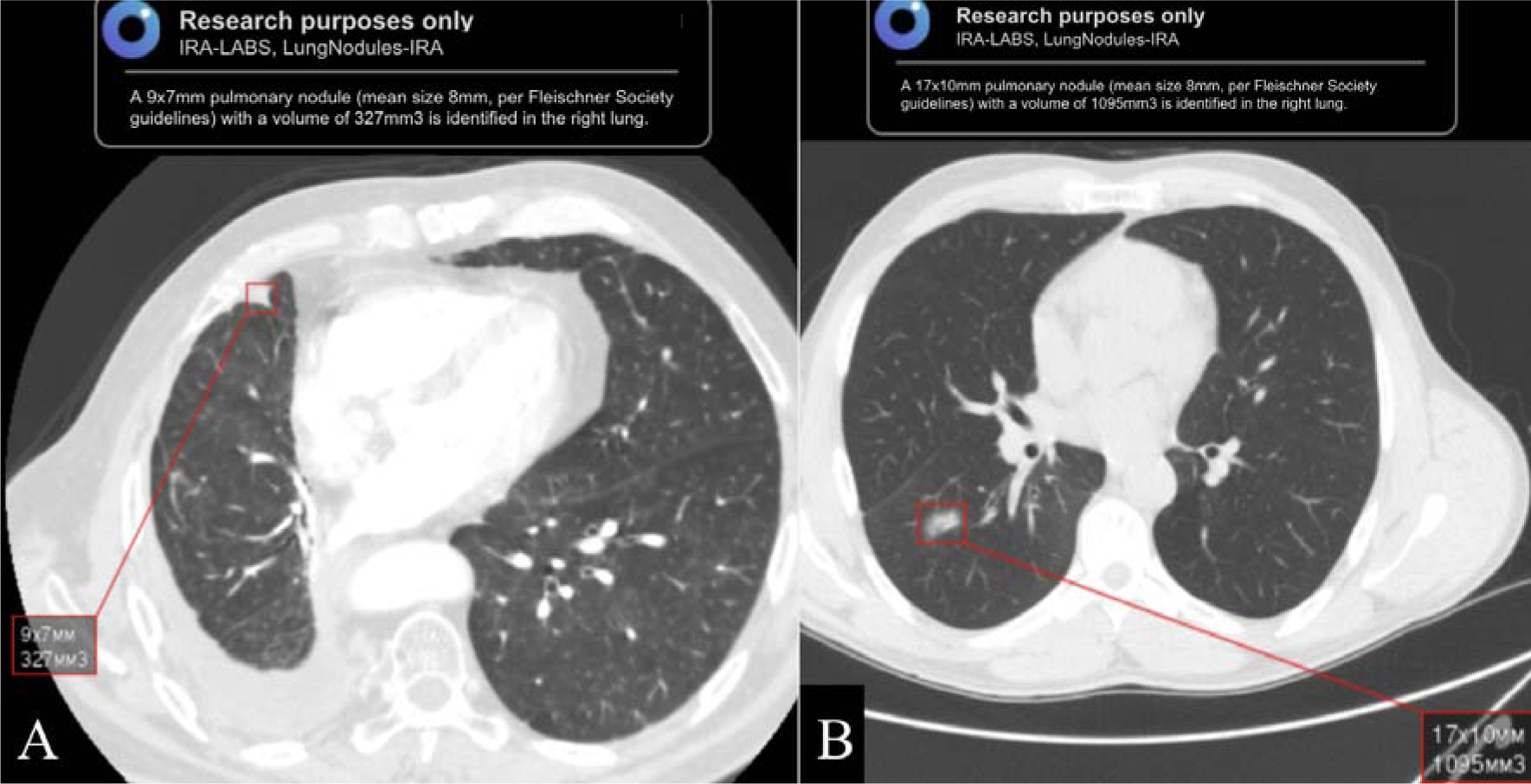
The following examples illustrate frequent false positives generated by the AI algorithm: A - fibrotic changes in the lung are erroneously identified as pulmonary nodule; B-area of lung tissue infiltration is mistakenly highlighted as pulmonary nodule.

### Evaluating cost-effectiveness

Evaluating the clinical effectiveness of an experiment is crucial, but it is also essential to predict the economic impact of AI implementation in the medical field.

To estimate potential labor costs, we used average data for calculations: the mean salary of a radiologist in Krasnoyarsk Krai is 96,900 rubles (or 1,066 USD, or 7,849 CNY) (Table 2). A radiologist typically processes approximately 400 CT studies per month, equivalent to 20 studies per day. Consequently, 10,500 studies from the initial sample would have required 26.25 months of a doctor’s work. In contrast, the AI independently analyzed 10,016 studies, saving 25.04 months of work, which would have cost the medical organization 2.43 million rubles in direct salary alone (excluding taxes and other payments) (or 27,421 USD, or 196,830 CNY).

**Table 2.** Comparing the economic impact of radiologist revision models with and without AI integration.

| **Parameters** | **Model #1**<br>*Review of all CT studies by radiologists without AI using* | **Model #2**<br>*The review process consists of two stages: initially, all CTs are assessed using AI, followed by radiologists examining only those CTs where AI has identified potential signs of lung cancer.* |
| --- | --- | --- |
| Initial number of CTs | 10,500 CTs |  |
| Number of CTs excluded from review by radiologists | not applicable | 10,016 CTs |
| Number of CTs reviewed by radiologists | 10,500 CTs | 484 CTs |
| The cost of radiologists' time for reviewing CTs (assuming a radiologist reviews 400 CT scans per month) | 26.25 months of work | 1.21 months of work |
| Cost of radiologist's work on review (assuming a radiologist's monthly salary of 96,900 rubles) | RUB 2,543,625 (or 27,980 USD, or 206,033 CNY) | RUB 117,249 (or 1,290 USD, or 9,497 CNY) |
| Time saved | 0 months of work | 25.04 months of work |
| Resources conserved (presuming AI implementation in the current project was costless) | RUB 0 | RUB 2,426,376 (or 26,690 USD, or 196,536 CNY) |

Another approach to assess the effectiveness of AI implementation is by examining the cost savings in treating patients at early stages of oncopathology compared to late stages. The application of AI algorithms enabled such retrospective analysis and identified 35 patients with early-stage LC for every 10,500 studies screened.

Estimating the average treatment cost for patients with stage I to II lung cancer, based on data from the Krasnoyarsk Krai Clinical Onco Dispensary in 2023, amounts to 352,000 rubles per patient (equivalent to approximately 3,872 USD or 28,512 CNY). Consequently, the total cost of early-stage care for the identified 35 patients would be 12,35 million rubles (or 135,811 USD or 1,000,066 CNY) (Table 3). In comparison, the estimated cost for patients with LC detected at later stages, with an average cost per patient of 587,000 rubles (or 6,465 USD or 47,606 CNY), is 20.57 million rubles (or 226,278 USD, or 1,666,229 CNY). Thus, the direct expected savings for the regional budget due to early-stage detection is 8,22 million rubles for every 10,500 individuals who underwent CT screening (or 90,466 USD or 666,162 CNY).

**Table 3.** Estimated economic impact of reducing the cost of treating patients with advanced lung cancer by conducting an AI project.

| **Parameters** | **Cost of treating a single patient** | **Cost of treatment for all early-detected patients**<br>**(n = 35)** |
| --- | --- | --- |
| Cost of treatment for stages I-II | RUB 352,757<br>(or 3,880 USD, or 28,573 CNY) | RUB 12,346,495<br>(or 135,811 USD, or 1,000,066 CNY) |
| The cost of treatment for stages III-IV<br>(combined treatment: surgery + chemoradiotherapy, excluding the cost of immunotherapy) | RUB 587,735<br>(or 6,465 USD, or 47,606 CNY) | RUB 20,570,725<br>(or 226,278 USD, or 1,666,229 CNY) |
| Cost-effectiveness from reduced costs of treating patients with advanced stages (excluding the cost of immunotherapy) | RUB 234,978<br>(or 2,585 USD, or 19,033 CNY) | RUB 8,224,230<br>(or 90,466 USD, or 666,162 CNY) |

A widely accepted method for evaluating the economic impact of healthcare interventions involves considering the years of life saved and their potential contribution to the gross regional product (GRP). The five-year survival rate for patients diagnosed with early-stage disease is approximately 90%. Consequently, this translates to a potential saving of 157,5 person-years of life for 35 persons. Using data from the Federal State Statistics Service (Rosstat) for the Krasnoyarsk Krai region, the estimated GRP is 1,1 million rubles (equivalent to approximately 12,100 USD or 89,100 CNY). This allows us to estimate the financial equivalent of the saved life years at 173,25 million rubles (equivalent to 1,905,750 USD or 14,033,250 CNY) (Table 4).

**Table 4.** Estimation of overall savings from AI when CT data are reused to detect evidence of pulmonary malignancy.

| **Parameters** | **Savings in the project** |  |
| --- | --- | --- |
|  | **Calculation for 1 year** | **Calculation for 5 years, taking into account years of life saved** |
| Reducing the radiologists' workload | RUB 2,426,376<br>(or 26,690 USD, or 196,536 CNY) |  |
| Decreased treatment expenses for late-stage patients, with the assumption of 35 early-stage patients | RUB 8,224,230<br>(or 90,466 USD, or 666,162 CNY) |  |
| The evaluation of the economic impact of life years saved on gross regional product over a five-year period | - | RUB 173,250,000<br>(or 1,905,750 USD, or 14,033,250 CNY) |
| Total economic effect | RUB 10,650,606<br>(or 117,157 USD, or 862,699 CNY) | RUB 183,900,606<br>(or 2,022,907 USD, or 14,895,949 CNY) |

Moreover, it is appropriate to consolidate the estimated impacts, as the concurrent benefits of early detection facilitated by AI applications have been observed. These benefits include time and labor savings for medical professionals, reduced treatment costs, and the potential contribution to regional gross product resulting from an increased five-year survival rate. Consequently, the cumulative effect can be approximated at 183,9 million rubles for 35 patients identified at an early stage per 10,500 CT examinations conducted over a five-year period (equivalent to 2,022,907 USD or 14,895,949 CNY).

## Discussion

In the retrospective analysis utilizing the AI algorithm, 355 CTs were chosen from a pool of 10,500 processed CT studies, after excluding defective scans. Upon review by specialists from the KKCOD, 252 CTs (2.4%) were classified into two groups: “patients with a high probability of LC” and “patients with insufficiently convincing signs of LC.” Among 100 histologically verified LC cases, 35 were in stages I-II and 65 were in stages III-IV. The direct expected savings for the regional budget due to early detection of patients are estimated to be 8,22 million rubles for every 10,500 individuals who underwent CT scans (or 90,466 USD, or 666,162 CNY). The total effect, which includes labor fund savings, reduction of treatment costs, and potential contribution to the regional gross product, is projected to be 183,9 million rubles (or 2,022,907 USD, or 14,895,949 CNY) over a five-year period.

However, the estimated number could be higher, as these calculations do not account for two groups of patients: first, those with depersonalization errors; second, those who received a postmortem diagnosis of malignant disease between the CT scan and the start of the retrospective study. If we model the enrollment of all 484 patients without depersonalization errors, considering the distribution of early stages as in the group of 355 known patients, the number of patients detected by AI in the early stages of lung cancer would be 43 patients. Consequently, the total economic benefit for these patients (2.43 million from revision + 10.105 million from treatment savings + 236.5 million from life years saved) would amount to 249.08 million rubles over 5 years (or 2,739,880 USD, or 20,175,480 Yuan).

The AI algorithm demonstrated reliable identification of nodular masses with high and intermediate likelihood of LC in the majority of cases, even amidst inflammatory changes due to COVID-19 infection. This underscores the efficacy of a hybrid methodology in the retrospective analysis of chest CTs.

It is important to note that in the Russian Federation, the percentage of LC cases identified at early stages (stage I-II) does not surpass 30% on average [9]. One contributing factor to this situation is the limited effectiveness of fluorographic examinations in detecting LC at early stages, which are conducted during routine health check-ups for the adult population. Identifying tumors larger than 1 cm, even with advanced radiological equipment, necessitates a high level of expertise from radiologists and a thorough evaluation of each patient’s individual risk factors, including genetic predisposition, occupational exposure, infectious diseases, and chronic conditions. Consequently, this method primarily detects late-stageLC, at which point radical treatment is no longer feasible, and the prognosis is generally unfavorable [9].

A crucial approach to addressing LC is through high-quality and timely screening. The most effective method for LC screening is low-dose computed tomography (LDCT), which has been shown to detect 3 to 4 times more focal masses compared to radiography, with the detected foci being half the size. The largest study on this subject, the National Lung Cancer Screening Trial (NLST) conducted in the United States, demonstrated that using LDCT as a screening method reduced LC mortality by 15-20% in comparison to lung radiography [9]. In 2015, LDCT was introduced as a pilot project for LC screening in Krasnoyarsk, targeting specific segments of the adult population [11]. The screening was conducted on individuals within the high-risk group, specifically men aged 50-64 years living in Krasnoyarsk with a pack/year index of 30 or more. As a result of implementing LDCT as a standard for LC screening between 2015 and 2017, the detection rate of LC reached 17.1 per 1000 examined patients. This detection rate was 30 times higher than that of lung malignancy detection achieved through programs utilizing fluorography or radiography, which had a rate of 0.57 per 1000 subjects [12]. In 2018, this approach was expanded throughout the Krasnoyarsk region, and by 2019, LDCT was incorporated into the standard medical examination protocol for certain adult population groups, with further expansion of the criteria for inclusion in the high-risk group (Fig. 7).

**Figure 7.**
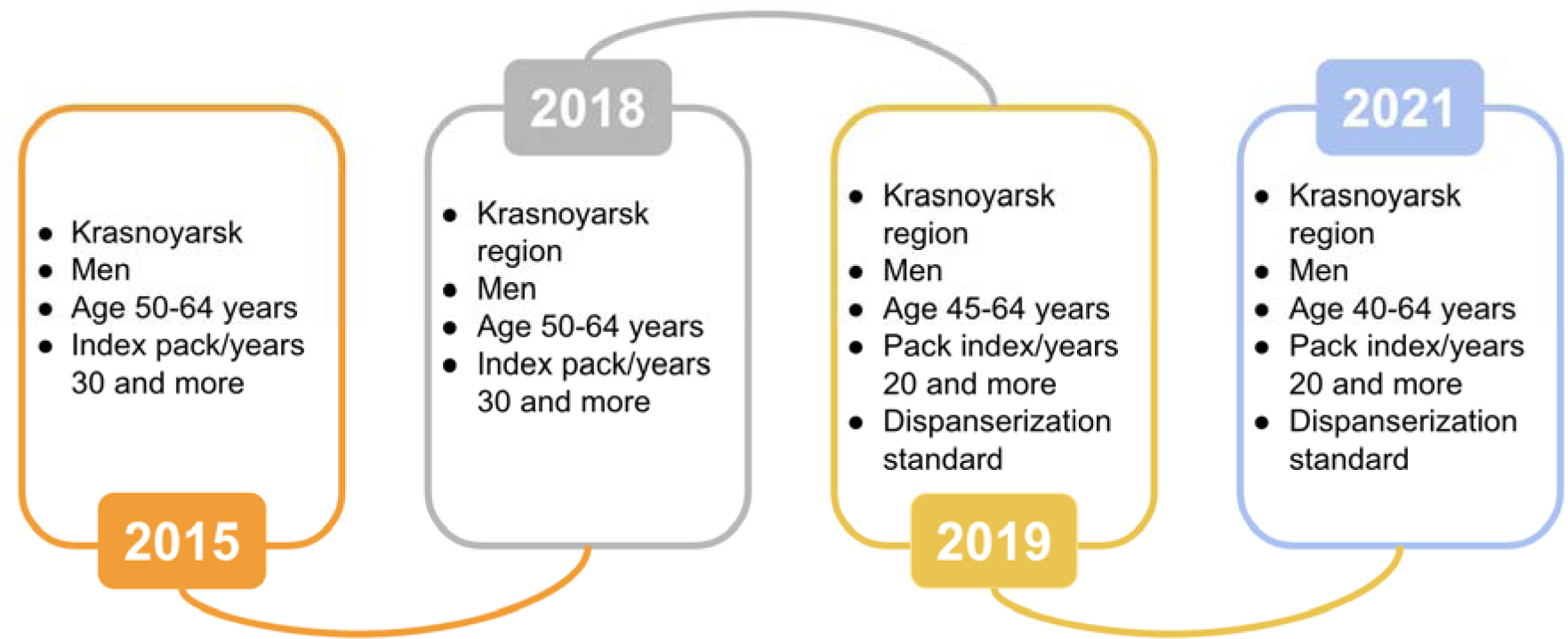
Progression of Lung Cancer Screening in the Krasnoyarsk Region, Russia.

Between 2015 and 2021, the implementation of LDCT for LC screening in Krasnoyarsk Krai, Russia, resulted in a significant increase in the early detection of lung cancer. The rate of early detection in Krasnoyarsk Krai rose from 22.1 per 100,000 population in 2015 to 34.2 per 100,000 population in 2021, representing a 54.8% increase. In comparison, the Russian Federation as a whole experienced an 8.4% increase in early detection rates during the same period, from 27.3 per 100,000 population in 2015 to 29.6 per 100,000 population in 2021. Furthermore, Krasnoyarsk Krai observed a 7.0% decrease in LC mortality rates between 2015 and 2021, from 48.7 per 100,000 population in 2015 to 45.3 per 100,000 population in 2021. This contrasts with the overall trend in the Russian Federation, where LC mortality rates increased by 13.2% during the same period, from 34.1 to 38.6 per 100,000 population [2].

Recent studies have demonstrated that AI algorithms can predict the risk of LC with a performance comparable to that of radiologists. AI-based comprehensive LC screening achieved state-of-the-art performance, with a ROC AUC of 94.4%. Furthermore, AI algorithms have been shown to improve radiologists’ diagnostic accuracy by reducing false positives by 11% and false negatives by 5% [11. A recent systematic review assessing the effectiveness of AI in the diagnosis and prognosis of lung malignancies revealed that radiologists utilizing AI achieved higher sensitivity and AUC, albeit with a slight decrease in specificity. In general, when predicting malignancy, radiologists using AI demonstrated higher sensitivity, specificity, and AUC [13].

In their study, Goncalves et al. conducted a SWOT (strengths, weaknesses, opportunities, threats) analysis to assess the strengths, weaknesses, opportunities, and threats associated with the application of AI in LC detection [14]. The authors identified several strengths, including the high accuracy of contemporary algorithms, the potential to alleviate the workload of physicians, and the reduction of diagnostic errors. However, they also emphasized the necessity for validation studies in real-world settings prior to clinical implementation as a weakness. Regarding threats, the study highlighted concerns related to confidentiality and data protection, as well as potential biases in the samples used for training AI algorithms. These issues underscore the need for careful consideration and ongoing research to ensure the safe and effective integration of AI in LC detection and diagnosis.

In our investigation, specialists detected false positives generated by the AI algorithm in 17% of all CTs where the AI recognized pulmonary nodules, totaling 60 CT scans. Unfortunately, these conclusions were based only on the opinions of the radiologists, without repeat CT scans, which may have influenced the results, reducing the results of the project.

Of the 104 CT scans identified by AI and confirmed by radiologists and invited for a repeat CT scan, only 78 (75%) people underwent a follow-up CT scan. Considering the significant time interval between CT and AI (up to 881 days), this indicator of patient involvement in verification is assessed as high.

A recent study conducted by Ziegelmayer S and colleagues assessed the cost-effectiveness of implementing an AI algorithm in the initial round of LC screening through the use of Markov simulation. The findings of the study indicate that incorporating AI assistance during the initial screening phase proves to be a cost-effective approach, with a maximum cost of 1,240 USD per patient, assuming a willingness to pay 100,000 USD for quality-adjusted life years (QALYs) [15]. QALYs are a measure of disease burden that combines both the quality and quantity of life lived, and are commonly used in economic evaluations of medical interventions. The proposed project, which involves reanalyzing CT data using AI to identify indications of LC, also demonstrates economic feasibility in both the short term and over a five-year period. The cost savings generated by this approach can be attributed to a significant decrease in the necessity for physician review of CTs, a reduction in the expenses associated with treating patients diagnosed with late-stage disease, and a future contribution to the regional economy through the preservation of life years.

In addition, this study examines the practical impact of harms from medical AI that partially replicate the harms found in lung cancer screening. In this project, harms from the use of AI:

- false positives that burden radiologists with having to revise CT images that did not need it (benign findings);
- unnecessary burden on radiology departments in the form of repeated invitations and CT examinations for those who, according to the results, did not have verified lung cancer;
- excessive burden on the health care system in the form of additional additional follow-up examinations and even surgical interventions for patients after CT scanning of the chest organs for those patents who, according to the results, did not have verified lung cancer;
- overdiagnosis to those patients for whom the established diagnosis will not cause their death, but will complicate the rest of their lives;
- the moral burden of not being able to invite for follow-up examinations those who have every reason to do so. These are those patients with findings suggested by AI, confirmed by physicians, patients still alive, not included in the Chancery Register, but for various reasons have not come for a follow-up chest CT scan;

In the current environment, we have no practical guidance on how to safely incorporate this technology into clinical workflows (16). But it is worth working to minimize these harms and conducting a comprehensive assessment of the balance of benefits and harms of planning for the practical application of medical AI.

### Limitations

This research presents several limitations.

- Firstly, the study was conducted retrospectively, which aligns with the stated purpose but may affect the design choice. The very long period of time between CT and the use of AI influenced the detection of a small number of newly diagnosed lung cancers. Future studies and recommendations should take this finding into account by limiting the retrospective data interval. The shorter the time between CT and AI, the higher the expected benefit (the number of newly diagnosed lung cancers).
- Secondly, each study was reviewed by a single radiologist, a decision made to conserve resources and expedite the review process for a large dataset. Not all patients with findings from AI underwent follow-up CT. Inviting patients based on AI analysis is a new challenge for healthcare organizations and could form the basis for the development of new recommendations.
- Thirdly, the study did not include data from patients whose pulmonary nodules, greater than 100 mm³, were not detected by the AI algorithm. This omission could potentially lead to an underestimation of the false negatives in the AI performance evaluation, a limitation necessitated by the resource constraints of the project.
- Fourthly, the AI was primarily designed to detect pulmonary nodules, not specifically LCs. Consequently, the findings detected by the AI encompassed a variety of conditions, not solely LCs. It is therefore recommended that a separate study be planned to assess the conversion from AI-suggested findings to confirmed LCs.
- Fifthly, a significant portion of patient losses during de-anonymization is primarily due to the technical unpreparedness of medical organizations for such projects. It is planned to discuss the creation of recommendations for conducting similar projects in order to reduce such errors.
- Finally, the cost-effectiveness estimation did not account for potential savings from providing immunotherapy to patients in advanced stages, nor did it consider direct and indirect costs.

## Conclusion

The application of AI in analyzing large volumes of chest CTs, performed for purposes unrelated to cancer screening, such as general screening or screening for other lung diseases like COVID-19, may enable the effortless and early identification of incidental pulmonary nodules that could otherwise go undetected. Considering the outcomes of the project in 100 LC detection, including new LC and economic potential assessment, the utilization of AI algorithms can be considered for broader adoption in large retrospective projects. Nonetheless, it remains imperative to persist in exploring the influence of AI on the initial diagnosis of pulmonary malignancies.

## Additional information Funding Source

This study was financed solely by the authors.

## Data Availability

All data produced in the present study are available upon reasonable request to the authors.

## List of abbreviations

AI: artificial intelligence
CT: computer tomography
GRP: gross regional product
LDCT: low dose computed tomography
LC: lung cancer
NLST: National Lung Screening Trial
KKCOD: Krasnoyarsk Krai Clinical Oncologic Dispensary named after A.I. Kryzhanovsky
QALYs: quality-adjusted life years
ROC AUC: receiver operating characteristic area under the curve
SWOT: strengths, weaknesses, opportunities, threats

## Acknowledgment

The authors would like to thank AstraZeneca for providing their expertise and scientific support.

## Authorship Compliance

In accordance with the International Committee of Medical Journal Editors (ICMJE) criteria, the authors confirm that their involvement in this study meets the international standards for authorship. All authors have made significant contributions to the study’s conceptualization, execution, and manuscript preparation. Furthermore, each author has reviewed and approved the final version of the manuscript prior to publication.

